# Telemedicine Utilization Trajectories and Sociodemographic Differences, 2019-2024

**DOI:** 10.1101/2025.03.26.25324648

**Authors:** Bingyu Zhang, Lu Li, Yiwen Lu, Jingchuan Guo, Jiang Bian, John B. Salmon, Robert L. Stetson, Michael A. Horst, Srinivas K. Sridhara, Mitchell D. Schnall, Kevin B. Mahoney, David A. Asch, Yong Chen

## Abstract

**IMPORTANCE:** Telemedicine usage surged during the COVID-19 pandemic, shaping how patients access healthcare services. Its sustained role in post-pandemic healthcare may uncover long-term trends and variations in utilization.

**OBJECTIVE:** To characterize telemedicine utilization patterns from 2019 to 2024 and identify patient characteristics associated with telemedicine use.

**DESIGN, SETTING, AND PARTICIPANTS:** This retrospective cohort study analyzed outpatient visits across five hospitals within the University of Pennsylvania Health System (Penn Medicine) from January 1, 2019, to September 30, 2024.

**MAIN OUTCOMES AND MEASURES:** The primary outcome was the proportion of visits conducted through telemedicine. Multivariable logistic regression models were employed to assess association between telemedicine use and patient characteristics including demographics, insurance type, patient portal use, and socioeconomic status.

**RESULTS:** The study included 46,149,734 visits among 2,248,341 patients. Following the declaration of the COVID-19 pandemic in March 2020, telemedicine surged from 1% to 17% of outpatient encounters by April 2020, stabilizing at 8-13% for the rest of the year. Usage declined in 2021 but remained at 4-6% from 2022 to 2024. In multivariable models, older adults were less likely to use telemedicine compared to those under 40 years (40-64 years: aOR, 0.67 [95% CI, 0.67-0.67]; ≥65 years: aOR, 0.46 [95% CI, 0.46-0.46]). Higher telemedicine use was observed among women (male: aOR, 0.91 [95% CI, 0.91-0.92]), unmarried individuals (aOR, 1.10, 95% CI, 1.10-1.11), patient portal users (aOR, 1.44 [95% CI, 1.43-1.45]), patients with fewer comorbidities (Charlson Comorbidity Index scores ≥3: aOR, 0.87 [95% CI, 0.87-0.88]), those living farther from the place of service (5-15 miles: aOR, 1.04 [95% CI, 1.04-1.04]; ≥15 miles: aOR, 1.44 [95% CI, 1.43-1.44]; reference: <5 miles), lower-income individuals (<$50,000: aOR, 1.06 [95% CI, 1.06-1.07]; ≥$100,000: aOR, 0.91 [95% CI, 0.91-0.92]; reference: $50,000-$100,000), and primary care compared to specialty care (aOR, 1.19 [95% CI, 1.18-1.20]). Return patients used telemedicine more than new patients (new: aOR, 0.47 [95% CI, 0.47-0.47]). Telemdicine use varied by race/ethnicity, with lower use among Non-Hispanic Black (aOR, 0.89 [95% CI, 0.88-0.89]), Hispanic (aOR, 0.95 [95% CI, 0.95-0.96]), and Asian (aOR, 0.83 [95% CI, 0.82-0.83]) patients compared to Non-Hispanic White patients. Patterns varied across visit types (e.g., diabetes, mental disorders, sleep disorders, heart failure, COPD, CAD, and GI disorders), though younger, female, and geographically distant patients consistently used telemedicine more. Non-Hispanic White patients with mental disorders exhibited disproportionately higher telemedicine use, underscoring racial/ethnic differences that persisted during and after the pandemic, likely influenced by differences in access and coverage.

**CONCLUSIONS AND RELEVANCE:** Telemedicine use is higher among tech-friendly populations, including, younger individuals, female, return patients, and those living farther from healthcare facilities. However, difference by age, socioeconomic status, and race/ethnicity persist, suggesting barriers in access, digital literacy, and coverage. Targeted policies are needed to ensure equitable telemedicine adoption and accessibility for all patients.

**Key Points:** *Question:* What has been the trajectory of telemedicine usage since the COVID-19 pandemic, and which sociodemographic factors are associated with its persistent use?

*Findings:* Telemedicine visits to the University of Pennsylvania Health System (Penn Medicine) surged with the onset of the COVID-19 pandemic in 2020. While telemedicine has since remained a significant component of outpatient care, its use has been uneven. Younger adults, women, patient portal users, unmarried individuals, return patients, and those with lower Charlson Comorbidity Index scores, were more likely to use telemedicine services. Moreover, racial and ethnic minorities, as well as socioeconomically disadvantaged populations, displayed varying patterns of telemedicine access, highlighting differences in utilization.

*Meaning:* Persistent differences in telemedicine use across primary care and specialty services highlight factors influencing its adoption, suggesting opportunities for targeted strategies to improve equity in digital healthcare access.

## Introduction

During the COVID-19 pandemic, most large health systems rapidly expanded their telemedicine capabilities to ensure continuity of patient care. Telemedicine was used for forward triage, automated patient screening, and integrating COVID-19 testing into care workflows.^1–4^ At the University of Pennsylvania Health System (Penn Medicine), this included tripling capacity for remote monitoring, incorporating virtual consultations at a newly opened inpatient facility, and leveraging digital tools to enhance patient safety and minimize in-hospital risks.^5^

Several studies have documented the rise in telemedicine encounters during the early and later phases of the pandemic, highlighting its adoption, peak utilization, and subsequent decline as in-person services resumed.^6–16^ However, most of these studies are limited to short-term trends, specific populations, or discrete time points within the pandemic, leaving gaps in understanding telemedicine’s long-term integration into routine care. Additionally, there remains a missed opportunity to comprehensively assess both telemedicine usage trajectories and patient characteristics.

This study aimed to address these gaps by leveraging longitudinal data from Penn Medicine, a large-scale health system with a patient population reflective of U.S. demographic diversity. By analyzing outpatient telemedicine use patterns from 2019 to 2024, this research examines pre-pandemic, pandemic, and post-pandemic trends, providing a unique perspective on the enduring role of telemedicine in routine care. The study also explored differences in patient characteristics between telemedicine and in-person visit users, investigated telemedicine usage across specific medical conditions, and identified populations where telemedicine remains underutilized. This comprehensive approach can not only uncover opportunities for more targeted telemedicine dissemination but also inform strategies to enhance equity and optimize its integration into post-pandemic healthcare.

## Methods

### Data and Study Population

This study used electronic health record (EHR) data from five hospitals within Penn Medicine, including the Hospital of the University of Pennsylvania (HUP), Penn Presbyterian Medical Center (PPMC), Pennsylvania Hospital (PAH), Chester County Hospital (CCH), and Penn Medicine Princeton Medical Center (PMC). Data were collected from January 1, 2019, to September 30, 2024.

The Penn Medicine EHR is a comprehensive, longitudinal data resource that supports clinical research across a wide spectrum of health conditions and treatments. It includes real-world data from inpatient and outpatient encounters across multiple hospitals and affiliated practices, covering over 6.5 million unique patients and more than 50 million clinical encounters. This study included data from one EHR instance across the system representing care provided in the Philadelphia region.

We included outpatient telemedicine and in-person encounters. Emergency department visits or inpatient visits were excluded. To ensure a meaningful comparison, we excluded visits primarily for lab tests, radiology, and telephones, focusing only on face-to-face visits. A complete list of included visit types is provided in the Supplementary Materials **eTable 1**.

### Primary Outcomes and Patient Characteristics

The primary outcome was telemedicine use at the visit level, defined as the proportion of visits conducted via telemedicine and modeled as a binary variable (telemedicine vs. in-person) in regression analyses. Patient characteristics considered in the analysis included: age (<40, 40-64, ≥65 years), sex (Male, Female), race/ethnicity (Hispanic, Non-Hispanic Black, Non-Hispanic White, Non-Hispanic Asian, Other/Unknown), insurance type (Commercial, Medicaid, Medicare, Self-Pay/Other), user of the patient portal (Yes, No), marital status (Married, Unmarried), median household income (estimated from zip code-level census data^17^, <$50,000, $50,000 to $100,000, ≥$100,000), encounter category (New patient visit, Return patient visit, Other/Unknown), Charlson Comorbidity Index scores^18^ (<3, ≥3), and distance from home to the place of service (<5 miles, 5-15 miles, ≥15 miles).

### Statistical Analysis

We examined the trajectory of telemedicine usage from 2019 to 2024. We categorized encounters by provider specialties and patient characteristics. We used multivariable logistic regression to estimate the association between these characteristics and the likelihood of a telemedicine visit.

To examine variations in telemedicine utilization for specific conditions, we conducted additional analyses focused on selected chronic conditions, including diabetes, mental disorders, sleep disorders, heart failure, chronic obstructive pulmonary disease (COPD), coronary artery disease (CAD), and gastrointestinal (GI) disorders. These conditions were chosen because they are common and might be well-suited for either in-person or telemedicine care. We repeated the above descriptive and regression models within these disease-specific subpopulations to investigate the trajectory and identify factors that may drive telemedicine usage uniquely in these groups. Two-sided P<0.05 was considered as statistically significant. All analyses were performed using R version 4.3.1.

## Results

### Cohort Identification

We identified 46,149,734 outpatient encounters among 2,248,341 patients within Penn Medicine between January 1, 2019, and September 30, 2024. Of these, 2,269,344 (4.9%) were completed via telemedicine, 43,880,390 (95.1%) occurred in person. Among all visits, 62.7% were women; 60.7% were Non-Hispanic White, 22.6% were Non-Hispanic Black, 4.8% were Hispanic, 4.7% were Non-Hispanic Asian; 78.7% were users of the patient portal.

Patients who completed telemedicine visits were more likely to be women, younger adults, patient portal users, return patients, unmarried, and with lower Charlson Comorbidity Index scores. A detailed comparison of baseline characteristics between telemedicine and in-person visits is summarized in **Table 1**.

**Table 1.** Baseline patient characteristics between telemedicine and in-person visits among outpatient encounters from 2019 to 2024.

|  | Telemedicine Visits<br>(N=2,269,344) | In-person Visits<br>(N=43,880,390) | Overall<br>(N=46,149,734) | Telemedicine<br>Utilization<br>Rate |
| --- | --- | --- | --- | --- |
| **Visit Year, no. (%)** |  |  |  |  |
| 2019 | 48763 (2.1) | 7421019 (16.9) | 7469782 (16.2) | 0.65% |
| 2020 | 700158 (30.9) | 6455362 (14.7) | 7155520 (15.5) | 9.78% |
| 2021 | 500002 (22.0) | 7505506 (17.1) | 8005508 (17.3) | 6.25% |
| 2022 | 395005 (17.4) | 7735083 (17.6) | 8130088 (17.6) | 4.86% |
| 2023 | 350273 (15.4) | 8165043 (18.6) | 8515316 (18.5) | 4.11% |
| 2024 | 275143 (12.1) | 6598377 (15.0) | 6873520 (14.9) | 4.00% |
| **Hospital, no. (%)** |  |  |  |  |
| CCH | 104373 (4.6) | 4122384 (9.4) | 4226757 (9.2) | 2.47% |
| HUP | 1311811 (57.8) | 20267793 (46.2) | 21579604 (46.8) | 6.08% |
| PMC | 293015 (12.9) | 5981315 (13.6) | 6274330 (13.6) | 4.67% |
| PAH | 277269 (12.2) | 6295894 (14.3) | 6573163 (14.2) | 4.22% |
| PPMC | 282876 (12.5) | 7213004 (16.4) | 7495880 (16.2) | 3.77% |
| **Age Range, no. (%)** |  |  |  |  |
| < 40 years | 830246 (36.6) | 10561066 (24.1) | 11391312 (24.7) | 7.29% |
| 40-64 years | 877811 (38.7) | 17008227 (38.8) | 17886038 (38.8) | 4.91% |
| ≥65 years | 561287 (24.7) | 16311097 (37.2) | 16872384 (36.6) | 3.33% |
| **Sex, no. (%)** |  |  |  |  |
| Female | 1510314 (66.6) | 27423206 (62.5) | 28933520 (62.7) | 5.22% |
| Male | 759030 (33.4) | 16457184 (37.5) | 17216214 (37.3) | 4.41% |
| **Race/Ethnicity, no. (%)** |  |  |  |  |
| Non-Hispanic White | 1417677 (62.5) | 26895385 (61.3) | 28313062 (61.4) | 5.01% |
| Non-Hispanic Black | 478386 (21.1) | 9948154 (22.7) | 10426540 (22.6) | 4.59% |
| Hispanic | 116809 (5.1) | 2083501 (4.7) | 2200310 (4.8) | 5.31% |
| Asian | 97128 (4.3) | 2082864 (4.7) | 2179992 (4.7) | 4.46% |
| Other/Unknown | 159344 (7.0) | 2870486 (6.5) | 3029830 (6.6) | 5.26% |
| **Insurance Plan, no. (%)** |  |  |  |  |
| Commercial | 1164593 (51.3) | 18903127 (43.1) | 20067720 (43.5) | 5.80% |
| Medicaid | 291334 (12.8) | 5084662 (11.6) | 5375996 (11.6) | 5.42% |
| Medicare | 595933 (26.3) | 15443489 (35.2) | 16039422 (34.8) | 3.72% |
| Self-Pay/Other | 217484 (9.6) | 4449112 (10.1) | 4666596 (10.1) | 4.66% |
| **MyPennMedicine User, no. (%)** |  |  |  |  |
| No | 302287 (13.3) | 9507494 (21.7) | 9809781 (21.3) | 3.08% |
| Yes | 1967057 (86.7) | 34372896 (78.3) | 36339953 (78.7) | 5.41% |
| **Marital Status, no. (%)** |  |  |  |  |
| Married | 1094117 (48.2) | 22795968 (52.0) | 23890085 (51.8) | 4.58% |
| Unmarried | 1175227 (51.8) | 21084422 (48.0) | 22259649 (48.2) | 5.28% |
| **Encounter Category, no. (%)** |  |  |  |  |
| Return Patient Visit | 1818743 (80.1) | 23247661 (53.0) | 25066404 (54.3) | 7.26% |
| New Patient Visit | 214514 (9.5) | 4309405 (9.8) | 4523919 (9.8) | 4.74% |
| Other/Unknown | 236087 (10.4) | 16323324 (37.2) | 16559411 (35.9) | 1.43% |
| **Median household income, no. (%)** |  |  |  |  |
| <\$50,000 | 426673 (18.8) | 8294546 (18.9) | 8721219 (18.9) | 4.89% |
| \$50,000 to \$100,000 | 725689 (32.0) | 14940258 (34.0) | 15665947 (33.9) | 5.13% |
| ≥\$100,000 | 1116982 (49.2) | 20645586 (47.0) | 21762568 (47.2) | 4.63% |
| **Charlson Comorbidity Index score, no. (%)** |  |  |  |  |
| <3 | 1407067 (62.0) | 23568167 (53.7) | 24975234 (54.1) | 5.63% |
| ≥3 | 862277 (38.0) | 20312223 (46.3) | 21174500 (45.9) | 4.07% |
| **Distance from Home to Place of Service, no. (%)** |  |  |  |  |
| < 5 miles | 773178 (34.1) | 16018615 (36.5) | 16791793 (36.4) | 4.60% |
| 5 to 15 miles | 773465 (34.1) | 16475670 (37.5) | 17249135 (37.4) | 4.48% |
| ≥15 miles | 722701 (31.8) | 11386105 (25.9) | 12108806 (26.2) | 5.97% |
| **Provider Specialty, no. (%)** |  |  |  |  |
| Primary Care | 247559 (10.9) | 2720825 (6.2) | 2968384 (6.4) | 8.34% |
| Specialty Care | 1950526 (86.0) | 25409781 (57.9) | 27360307 (59.3) | 7.13% |
| Unknown | 71259 (3.1) | 15749784 (35.9) | 15821043 (34.3) | 0.45% |

### Telemedicine Utilization Trajectories

Figure 1 illustrates the monthly number of telemedicine and in-person visits from 2019 to 2024. From 2019 to early 2020, telemedicine usage was minimal (around 1%). With the onset of COVID-19 in March 2020, telemedicine visits surged from 1% to 17% by April and fluctuated between 8-13% for the rest of the year. Utilization declined in 2021 and stabilized around 4-6% from 2022 onward, suggesting a continued but reduced role for telemedicine as in-person visits regained prominence.

**Figure 1.**
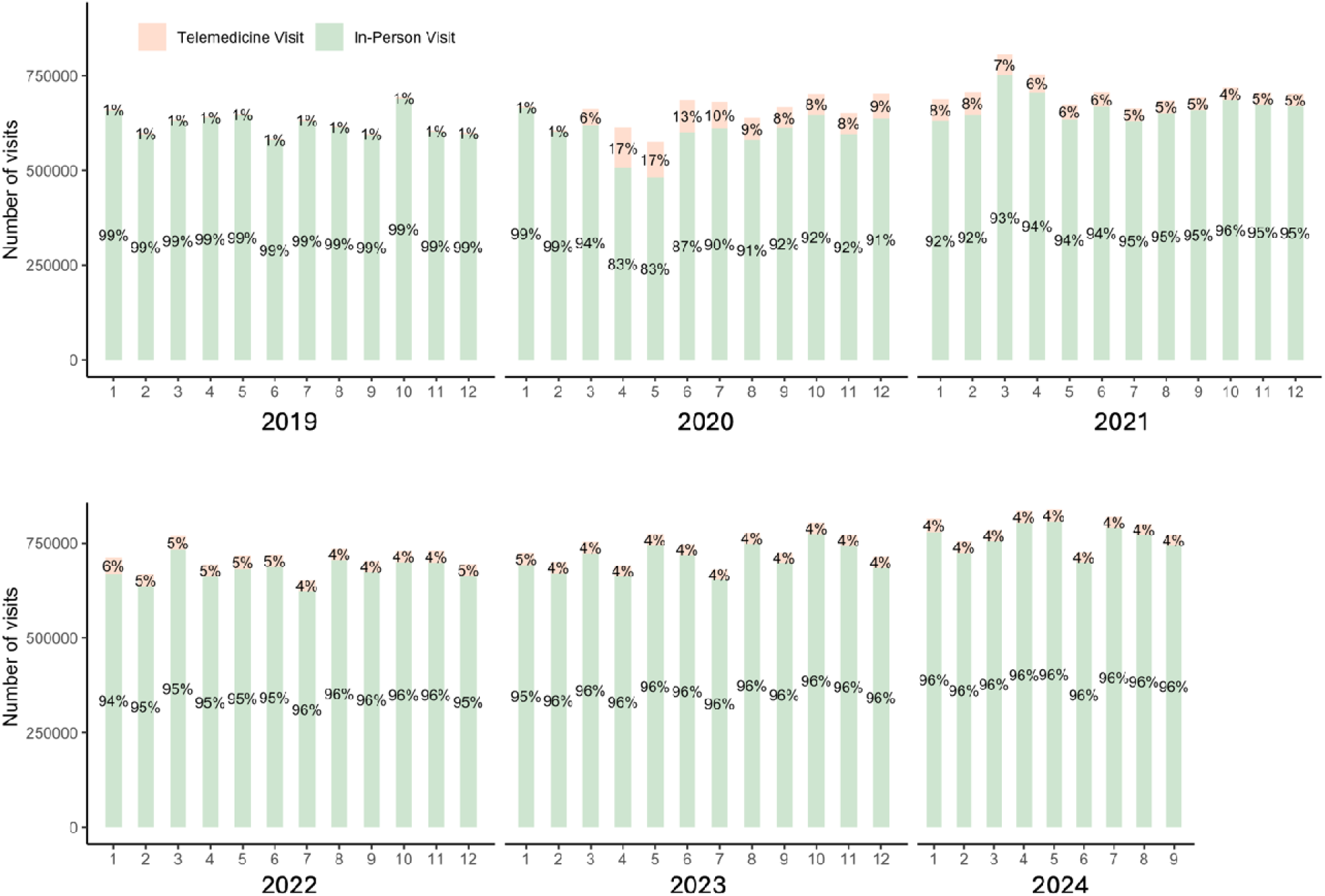
Telemedicine utilization rate by calendar time, monthly from 2019 to 2024.

Figure 2a shows the telemedicine utilization rates categorized by provider specialties. Sleep medicine, and psychiatry & neurology saw the highest telemedicine use, peaking at 20-35% in 2021 and remaining relatively high into 2024. Family medicine, internal medicine, surgery, and obstetrics & gynecology peaked at around 10-15% and declined to 5%. Nursing services had the lowest telemedicine rates, underscoring the continued need for in-person care in those fields. Only specialties with high prevalence were reported; the full list is available in the Supplementary Materials **eTable 2**.

**Figure 2.**
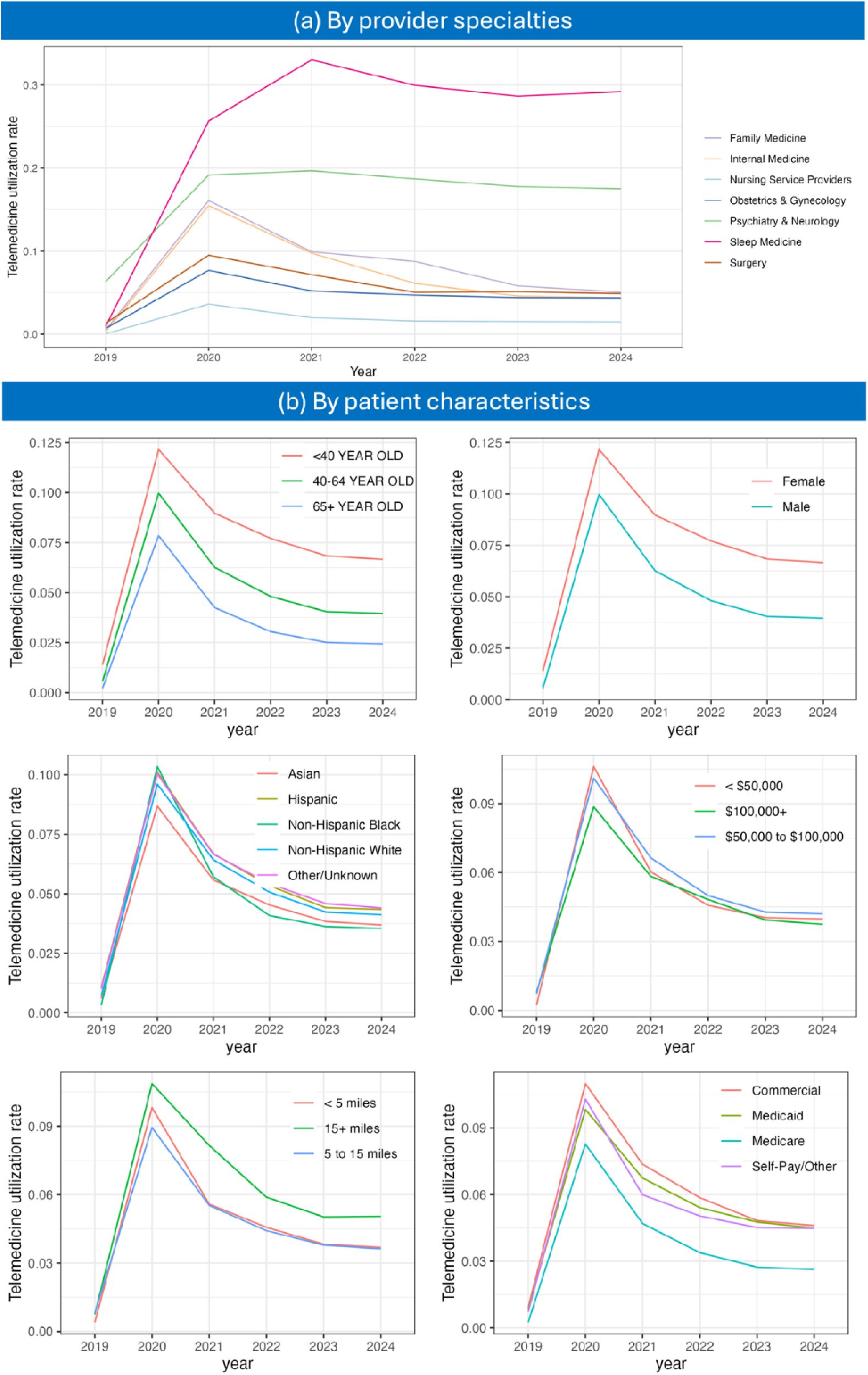
Trajectories of telemedicine visit rates categorized by provider specialties and patient characteristics.

Figure 2b illustrates telemedicine utilization trajectories stratified by various patient characteristics. Younger individuals (<40 years) had the highest telemedicine adoption rates, while older adults (65+ years) consistently reported lower utilization. Females demonstrated higher telemedicine usage across all time points compared to males. Patients with household incomes exceeding $100,000 had a lower rate of telemedicine use. Non-Hispanic White patients reported slightly greater telemedicine usage than other racial and ethnic groups. Patients residing more than 15 miles from healthcare facilities showed higher telemedicine adoption rates than those living closer. Patients with commercial insurance had the highest telemedicine usage throughout the study period, whereas those with Medicare demonstrated lower rates.

### Factors Associated with Telemedicine Use

Compared with patients younger than 40 years, older patients were less likely to receive care via telemedicine (40-64 years: aOR, 0.67 [95% CI, 0.67-0.67]; ≥65 years: aOR, 0.46 [95% CI, 0.46-0.46]). Compared with Non-Hispanic White patients, Asian (aOR, 0.83, 95% CI, 0.82-0.83), Hispanic (aOR, 0.95, 95% CI, 0.95-0.96), and Non-Hispanic Black (aOR, 0.89, 95% CI, 0.88- 0.89) patients were associated with less telemedicine use. Male (aOR: 0.91, 95% CI, 0.91-0.92) and new patient visits (aOR: 0.47, 95% CI, 0.47-0.47) were associated with less telemedicine use. Unmarried patients (aOR, 1.10, 95% CI, 1.10-1.11) and patient portal users (aOR, 1.44, 95% CI, 1.43-1.45) were more likely to use telemedicine. Patients with higher Charlson Comorbidity Index scores (aOR, 0.87 [95% CI, 0.87-0.88]) were less likely to use telemedicine. A median household income greater than $100,000 was associated with slightly less telemedicine use compared with an income of $50,000 to $100,000 (aOR, 0.91, 95% CI, 0.91-0.92). Longer distance from home to the place of service is associated with more telemedicine usage (5 to 15 miles: aOR, 1.04, 95% CI, 1.04-1.04; ≥15 miles: aOR, 1.44, 95% CI, 1.43-1.44). Visits with a primary care provider had higher telemedicine utilization compared to specialty care (aOR, 1.19, 95% CI, 1.18-1.20). **Table 2** summarizes the results from the multivariable logistic regression model.

**Table 2.**
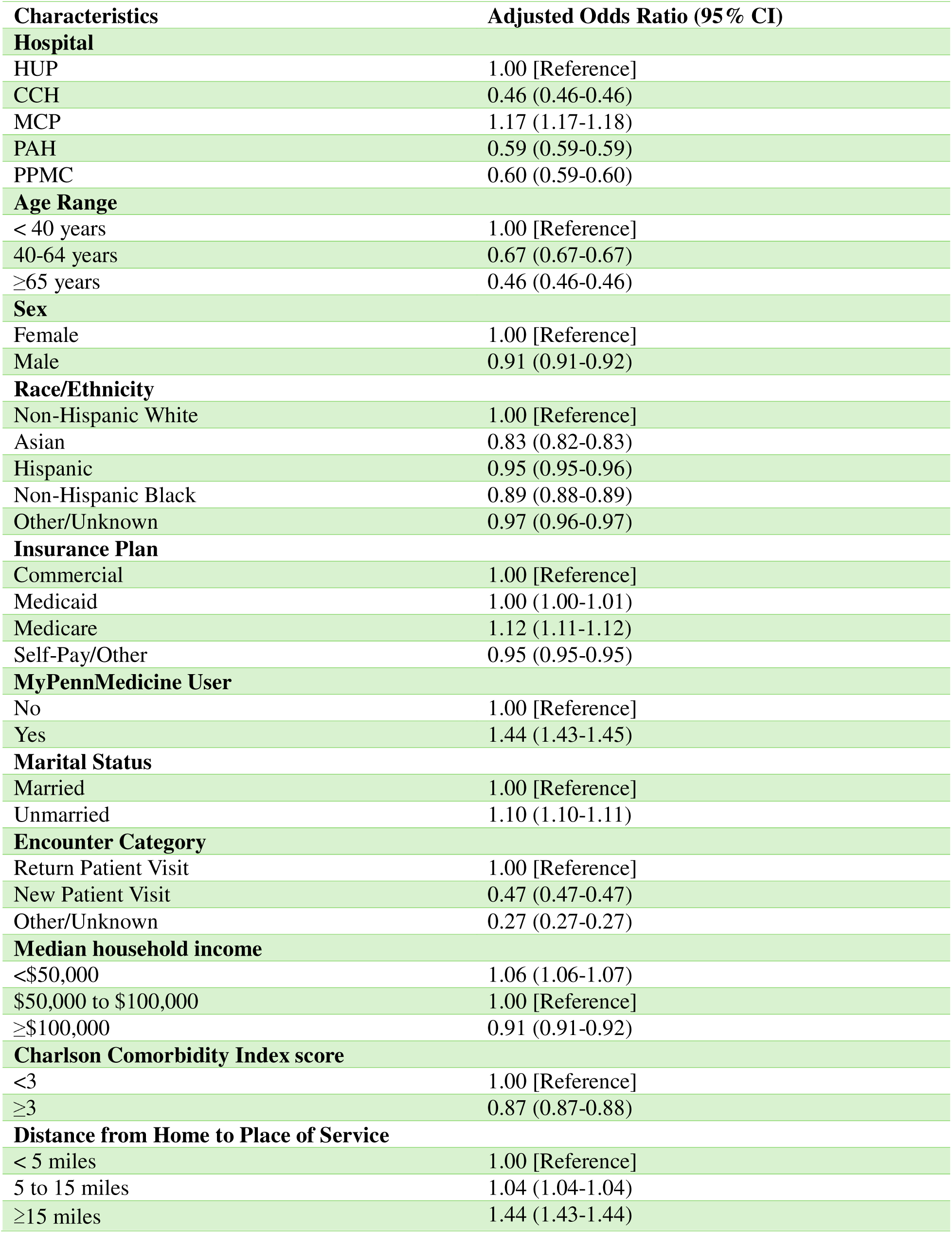
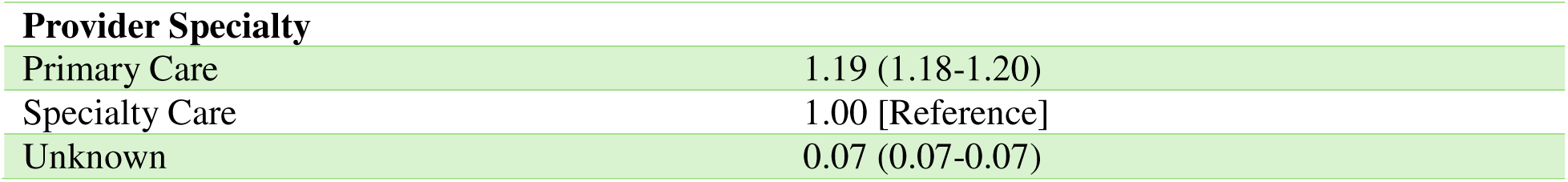
Multivariable logistic regression model associations with telemedicine and in-person visits among outpatient encounters from 2019 to 2024.

| Characteristics | Adjusted Odds Ratio (95% CI) |
| --- | --- |
| **Hospital** |  |
| HUP | 1.00 [Reference] |
| CCH | 0.46 (0.46-0.46) |
| MCP | 1.17 (1.17-1.18) |
| PAH | 0.59 (0.59-0.59) |
| PPMC | 0.60 (0.59-0.60) |
| **Age Range** |  |
| < 40 years | 1.00 [Reference] |
| 40-64 years | 0.67 (0.67-0.67) |
| ≥65 years | 0.46 (0.46-0.46) |
| **Sex** |  |
| Female | 1.00 [Reference] |
| Male | 0.91 (0.91-0.92) |
| **Race/Ethnicity** |  |
| Non-Hispanic White | 1.00 [Reference] |
| Asian | 0.83 (0.82-0.83) |
| Hispanic | 0.95 (0.95-0.96) |
| Non-Hispanic Black | 0.89 (0.88-0.89) |
| Other/Unknown | 0.97 (0.96-0.97) |
| **Insurance Plan** |  |
| Commercial | 1.00 [Reference] |
| Medicaid | 1.00 (1.00-1.01) |
| Medicare | 1.12 (1.11-1.12) |
| Self-Pay/Other | 0.95 (0.95-0.95) |
| **MyPennMedicine User** |  |
| No | 1.00 [Reference] |
| Yes | 1.44 (1.43-1.45) |
| **Marital Status** |  |
| Married | 1.00 [Reference] |
| Unmarried | 1.10 (1.10-1.11) |
| **Encounter Category** |  |
| Return Patient Visit | 1.00 [Reference] |
| New Patient Visit | 0.47 (0.47-0.47) |
| Other/Unknown | 0.27 (0.27-0.27) |
| **Median household income** |  |
| <\$50,000 | 1.06 (1.06-1.07) |
| \$50,000 to \$100,000 | 1.00 [Reference] |
| ≥\$100,000 | 0.91 (0.91-0.92) |
| **Charlson Comorbidity Index score** |  |
| <3 | 1.00 [Reference] |
| ≥3 | 0.87 (0.87-0.88) |
| **Distance from Home to Place of Service** |  |
| < 5 miles | 1.00 [Reference] |
| 5 to 15 miles | 1.04 (1.04-1.04) |
| ≥15 miles | 1.44 (1.43-1.44) |

| **Provider Specialty** |  |
| --- | --- |
| Primary Care | 1.19 (1.18-1.20) |
| Specialty Care | 1.00 [Reference] |
| Unknown | 0.07 (0.07-0.07) |

### Visits categorized by Different Health Conditions

Figure 3 illustrates detailed trajectories of patient characteristics for visits of various health conditions, including diabetes, mental, behavioral, and neurodevelopmental disorders (MBD), sleep disorders, heart failure, chronic obstructive pulmonary disease (COPD), coronary artery disease (CAD), and gastrointestinal (GI) disorders.

**Figure 3.**
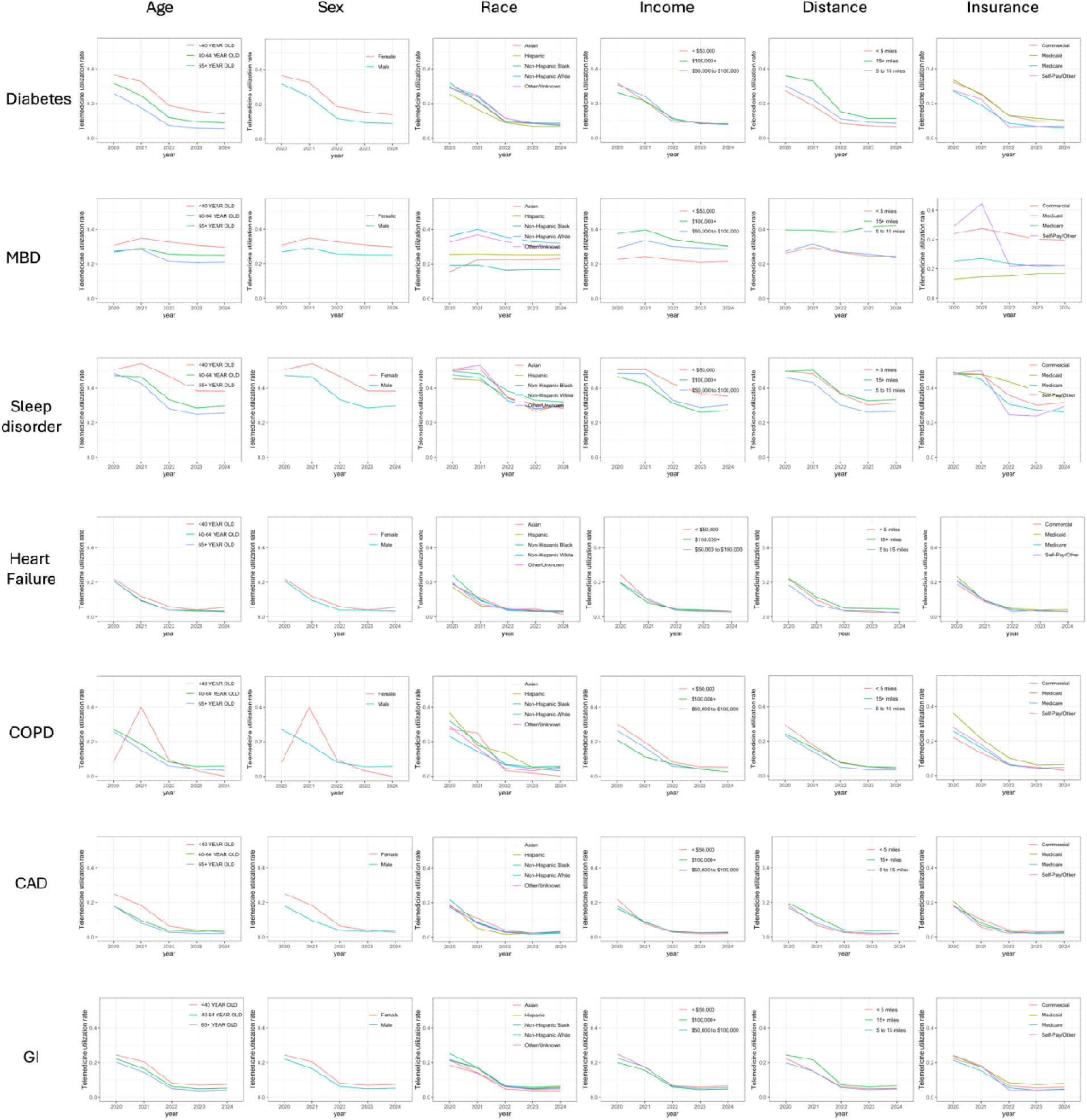
Trajectories of characteristics for visits of various health conditions, including diabetes, mental, behavioral, and neurodevelopmental disorders (MBD), sleep disorders, heart failure, chronic obstructive pulmonary disease (COPD), coronary artery disease (CAD), and gastrointestinal (GI) disorders.

Across these diagnoses, the trends in age, sex, and distance from home to the place of service remain consistent. Younger patients, female patients, and patients living further from the place of service are more likely to use telemedicine. Utilization rates for these conditions declined after 2021 but remained above pre-pandemic levels, likely reflecting the convenience of remote monitoring and follow-ups for chronic conditions that require routine management.

Telemedicine utilization for patients with diabetes peaked during the pandemic, particularly among younger patients. Sleep disorders demonstrated sustained high telemedicine utilization post-pandemic. For patients with heart failure and COPD, telemedicine use was more modest. Both CAD and GI disorders followed similar patterns of low-to-moderate telemedicine use.

In addition, a specific trend observed in MBD-related visits shows a higher proportion of telemedicine usage among Non-Hispanic White patients. This finding suggests potential differences in telemedicine utilization across racial/ethnic groups for MBD diagnoses. Furthermore, in 2020 and 2021, a greater percentage of MBD-related visits were with self-pay or other insurance types, though the percentage declined after 2022. This trend may be attributed to policy changes expanding coverage for MBD-related telemedicine visits under commercial insurance plans.

The multivariable regression results for visits within each health condition are available in Supplementary Materials **eTables 3-9**.

## Discussion

This study highlights the trends in telemedicine and in-person visits from Penn Medicine from 2019 to 2024, revealing significant shifts in care delivery during the COVID-19 pandemic. Our findings show a rapid surge in telemedicine use during the early months of the pandemic, followed by stabilization at lower but sustained levels as in-person care resumed. The initial spike reflects the healthcare system’s swift adaptation to pandemic restrictions, while the sustained utilization suggests that telemedicine has become a complementary mode of care delivery, particularly for follow-up visits and consultations that do not require physical examinations.

While these overall observations are not novel, our multivariable analysis highlighted key demographic and socioeconomic differences within these known telemedicine trends. Younger patients (<40 years) consistently exhibited higher telemedicine adoption across conditions, potentially reflecting greater comfort with technology. Conversely, older adults (65+ years) showed lower utilization rates, possibly due to barriers such as technology access or digital literacy.^19^ Additionally, female patients, individuals with lower household incomes, and patient portal users were more likely to use telemedicine.

The breakdown by specialty also aligns with findings from prior research, which reported sustained telemedicine use in psychiatry and primary care but lower adoption in procedural specialties such as surgery. These patterns underscore the importance of tailoring telemedicine strategies to the needs of specific patient populations and specialties. For example, incorporating hybrid care models that balance telemedicine with in-person visits could address the needs of patients with chronic conditions requiring both routine monitoring and hands-on evaluations.^20^

Beyond patient and specialty-level factors, systematic elements within Penn Medicine also influenced telemedicine adoption. Not all outpatient visits had equal opportunity for telehealth encounters, as internal scheduling algorithms directed certain visit types toward in-person care, even for routine chronic condition follow-ups. These algorithms, which evolved throughout the study period, likely contributed to variations in telemedicine utilization and may have created an upper limit on the number of visits eligible for telehealth. Additionally, provider and clinic-level biases played a role, as some providers actively embraced telehealth, while others remained resistant, even for visit types well-suited for remote care. This aligns with our findings that younger patients were more likely to use telemedicine—younger providers may also be more inclined to offer telehealth services to their patients, further reinforcing these trends. Understanding these structural and provider-driven influences is crucial for future telemedicine optimization.

The condition-specific analysis revealed that telemedicine use varied by the clinical nature of each condition. Mental health and sleep disorders maintained consistently high utilization rates, reflecting their suitability for remote care delivery. Conversely, conditions such as heart failure and COPD, which often require physical assessments and complex management, showed moderate telemedicine adoption, with rates declining post-pandemic. Chronic conditions like diabetes sustained use for routine monitoring and follow-ups, whereas procedural or diagnostic- heavy conditions, such as CAD and GI disorders, consistently relied more on in-person care. These variations emphasize the need to align telemedicine practices with the specific demands of different clinical conditions.

Limitations of this study include that the data are specific to Penn Medicine and may not generalize to other healthcare settings with different patient populations and telemedicine infrastructures. Also, while we analyzed trends over a multi-year period, telemedicine adoption was influenced not only by patient preference but also by institutional scheduling rules and provider-driven biases, which were not fully captured in our models. Future research should explore the impact of these system-level constraints on telemedicine availability and uptake.

## Conclusion

The COVID-19 pandemic catalyzed a rapid expansion of telemedicine, fundamentally reshaping care delivery. The sustained high utilization for mental health, sleep disorders, and chronic disease management highlights the potential for telemedicine to improve access and convenience for these populations.

However, differences in telemedicine adoption, particularly among older adults, certain racial/ethnic groups, and those with public insurance, emphasize the need for targeted interventions to promote equitable access. Future research should focus on optimizing hybrid care models, evaluating long-term patient outcomes, and understanding the impact of policy changes on telemedicine utilization.

## Data Availability

All data produced in the present study are available upon request

## Disclosures Funding

We acknowledge the start-up funding from University of Pennsylvania Health System.

## Conflicts of Interest

Dr. David A. Asch is a partner and part owner of VAL Health and serves on the advisory boards of Thrive Global and Morpheus. Dr. Yong Chen reported receiving personal fees from Merck & Co., Inc. outside the submitted work. All other co-authors have no conflicts of interest to report.

## Notes

### Author Declarations

Ethics committee/IRB of University of Pennsylvania gave ethnical approval for this work

