## Supplemental Materials for "Telemedicine Utilization Trajectories and Sociodemographic Differences, 2019-2024"

**Supplementary Materials to “Telemedicine Utilization Trajectories and Sociodemographic Differences, 2019-2024”**

### eTable 1. Face-to-face list of encounter types

| **Encounter Type** | **Encounter Type Name** |
| --- | --- |
| 2 | Walk-In |
| 3 | Hospital Encounter |
| 11 | Research Encounter |
| 50 | Appointment |
| 76 | Telemedicine |
| 81 | Ophth Exam |
| 91 | Home Care Visit |
| 101 | Office Visit |
| 108 | Immunization |
| 120 | Endoscopy |
| 200 | Allied Health Visit |
| 436 | PT/OT/ST |
| 438 | OBGYN Visit |
| 441 | In Home Primary Care Encounter |
| 449 | PEC Visit |
| 450 | OBGYN SURGERY |
| 452 | Out of Office Visit |
| 600 | General Surgery |
| 604 | Anticoagulation Therapy |
| 610 | Infusion Visit |
| 1003 | Procedure Visit |
| 2523 | Hospice F2F Visit |
| 32002 | Care Management |
| 32003 | Virtual Visit |
| 32006 | Psych Office Visit |
| 32009 | Psych Allied Health |
| 32012 | Procedure |
| 32013 | Allied Health Visit (Non-Chargeable) |
| 32030 | Telehealth |
| 32032 | Psych Allied Health (Non-Chargeable) |
| 304113 | Palliative Home Care Visit |
| 304114 | CCBH Scheduled |
| 304117 | Telemedicine (Non-Chargeable) |
| 1650000002 | TXP Evaluation |

### eTable 2. Complete list of provider specialties

The provider specialties were categorized according to <https://taxonomy.nucc.org/>.

**Provider Specialty Clusters**

Anesthesiology

Behavioral Health & Social Service Providers

Chiropractic Providers

Critical Care

Dental Providers

Dermatology

Dietary & Nutritional Service Providers

Emergency Medicine

Eye and Vision Services Providers

Family Medicine

Hospice and Palliative Medicine

Hospitalist

Hospitals

Internal Medicine

Medical Genetics

Nuclear Medicine

Nursing Service Providers

Obstetrics & Gynecology

Ophthalmology

Other Service Providers

Pathology

Pediatrics

Pharmacy Service Providers

Physical Medicine & Rehabilitation

Physician Assistants & Advanced Practice Nursing Providers

Podiatric Medicine & Surgery Service Providers

Preventive Medicine

Psychiatry & Neurology

Radiology

Residential Treatment Facilities

Respiratory, Developmental, Rehabilitative and Restorative Service Providers

Sleep Medicine

Speech, Language and Hearing Service Providers

Sports Medicine

Surgery

Urology

### eTable 3. Adjusted odds ratios for diabetes visits

| Characteristics | Adjusted Odds Ratio (95% CI) |
| --- | --- |
| Hospital |  |
| HUP | 1.00 [Reference] |
| CCH | 0.18 (0.17-0.2) |
| MCP | 0.49 (0.47-0.51) |
| PAH | 0.82 (0.79-0.86) |
| PPMC | 0.88 (0.85-0.9) |
| Age Range |  |
| < 40 years | 1.00 [Reference] |
| 40-64 years | 0.7 (0.68-0.73) |
| ≥65 years | 0.45 (0.43-0.47) |
| Sex |  |
| Female | 1.00 [Reference] |
| Male | 0.83 (0.81-0.85) |
| Race/Ethnicity |  |
| Non-Hispanic White | 1.00 [Reference] |
| Asian | 0.92 (0.88-0.97) |
| Hispanic | 0.92 (0.87-0.97) |
| Non-Hispanic Black | 0.95 (0.92-0.98) |
| Other/Unknown | 0.9 (0.86-0.95) |
| Insurance Plan |  |
| Commercial | 1.00 [Reference] |
| Medicaid | 1.16 (1.12-1.2) |
| Medicare | 1.16 (1.12-1.2) |
| Self-Pay/Other | 0.68 (0.64-0.71) |
| MyPennMedicine User |  |
| No | 1.00 [Reference] |
| Yes | 1.48 (1.42-1.53) |
| Marital Status |  |
| Married | 1.00 [Reference] |
| Unmarried | 0.97 (0.95-1) |
| Encounter Category |  |
| Return Patient Visit | 1.00 [Reference] |
| New Patient Visit | 0.47 (0.45-0.5) |
| Other/Unknown | - |
| Median household income |  |
| <$50,000 | 1.14 (1.1-1.18) |
| $50,000 to $100,000 | 1.00 [Reference] |
| ≥$100,000 | 1.05 (1.03-1.09) |
| Charlson Comorbidity Index score |  |
| <3 | 1.00 [Reference] |
| ≥3 | 0.84 (0.81-0.86) |
| Distance from Home to Place of Service |  |
| < 5 miles | 1.00 [Reference] |
| 5 to 15 miles | 1.35 (1.31-1.38) |
| ≥15 miles | 1.8 (1.75-1.86) |

### eTable 4. Adjusted odds ratios for MBD visits

| Characteristics | Adjusted Odds Ratio (95% CI) |
| --- | --- |
| Hospital |  |
| HUP | 1.00 [Reference] |
| CCH | 0.12 (0.12-0.13) |
| MCP | 0.06 (0.06-0.06) |
| PAH | 0.06 (0.06-0.06) |
| PPMC | 0.18 (0.17-0.18) |
| Age Range |  |
| < 40 years | 1.00 [Reference] |
| 40-64 years | 0.92 (0.9-0.93) |
| ≥65 years | 0.8 (0.78-0.82) |
| Sex |  |
| Female | 1.00 [Reference] |
| Male | 0.91 (0.9-0.92) |
| Race/Ethnicity |  |
| Non-Hispanic White | 1.00 [Reference] |
| Asian | 1 (0.97-1.02) |
| Hispanic | 1 (0.98-1.02) |
| Non-Hispanic Black | 0.82 (0.81-0.83) |
| Other/Unknown | 1.14 (1.12-1.16) |
| Insurance Plan |  |
| Commercial | 1.00 [Reference] |
| Medicaid | 0.74 (0.73-0.75) |
| Medicare | 0.81 (0.79-0.82) |
| Self-Pay/Other | 0.92 (0.91-0.94) |
| MyPennMedicine User |  |
| No | 1.00 [Reference] |
| Yes | 3.4 (3.33-3.46) |
| Marital Status |  |
| Married |  |
| Unmarried | 0.93 (0.92-0.94) |
| Encounter Category |  |
| Return Patient Visit | 1.00 [Reference] |
| New Patient Visit | 0.52 (0.51-0.53) |
| Other/Unknown | - |
| Median household income |  |
| <$50,000 | 1.03 (1.02-1.05) |
| $50,000 to $100,000 | 1.00 [Reference] |
| ≥$100,000 | 0.88 (0.86-0.89) |
| Charlson Comorbidity Index score |  |
| <3 | 1.00 [Reference] |
| ≥3 | 0.79 (0.77-0.8) |
| Distance from Home to Place of Service |  |
| < 5 miles | 1.00 [Reference] |
| 5 to 15 miles | 0.98 (0.96-0.99) |
| ≥15 miles | 1.37 (1.35-1.39) |

### eTable 5. Adjusted odds ratios for sleep awake disorder visits

| Characteristics | Adjusted Odds Ratio (95% CI) |
| --- | --- |
| Hospital |  |
| HUP | 1.00 [Reference] |
| CCH | 0.21 (0.2-0.22) |
| MCP | 0.26 (0.24-0.27) |
| PAH | 0.78 (0.76-0.81) |
| PPMC | 0.53 (0.51-0.55) |
| Age Range |  |
| < 40 years | 1.00 [Reference] |
| 40-64 years | 0.72 (0.7-0.74) |
| ≥65 years | 0.63 (0.6-0.65) |
| Sex |  |
| Female | 1.00 [Reference] |
| Male | 0.8 (0.78-0.81) |
| Race/Ethnicity |  |
| Non-Hispanic White | 1.00 [Reference] |
| Asian | 1.11 (1.06-1.17) |
| Hispanic | 1.02 (0.97-1.07) |
| Non-Hispanic Black | 1.06 (1.02-1.09) |
| Other/Unknown | 1.03 (0.99-1.08) |
| Insurance Plan |  |
| Commercial | 1.00 [Reference] |
| Medicaid | 1.12 (1.08-1.16) |
| Medicare | 1.09 (1.06-1.13) |
| Self-Pay/Other | 0.94 (0.9-0.97) |
| MyPennMedicine User |  |
| No | 1.00 [Reference] |
| Yes | 2.34 (2.25-2.43) |
| Marital Status |  |
| Married | 1.00 [Reference] |
| Unmarried | 1.06 (1.04-1.08) |
| Encounter Category |  |
| Return Patient Visit | 1.00 [Reference] |
| New Patient Visit | 0.26 (0.26-0.27) |
| Other/Unknown | - |
| Median household income |  |
| <$50,000 | 1.24 (1.2-1.28) |
| $50,000 to $100,000 | 1.00 [Reference] |
| ≥$100,000 | 1.04 (1.02-1.07) |
| Charlson Comorbidity Index score |  |
| <3 | 1.00 [Reference] |
| ≥3 | 0.8 (0.78-0.82) |
| Distance from Home to Place of Service |  |
| < 5 miles | 1.00 [Reference] |
| 5 to 15 miles | 1.01 (0.99-1.04) |
| ≥15 miles | 1.35 (1.31-1.39) |

### eTable 6. Adjusted odds ratios for heart failure visits

| Characteristics | Adjusted Odds Ratio (95% CI) |
| --- | --- |
| Hospital |  |
| HUP | 1.00 [Reference] |
| CCH | 0.32 (0.24-0.42) |
| MCP | 0.33 (0.28-0.4) |
| PAH | 1.02 (0.92-1.14) |
| PPMC | 0.76 (0.71-0.83) |
| Age Range |  |
| < 40 years | 1.00 [Reference] |
| 40-64 years | 0.93 (0.81-1.06) |
| ≥65 years | 0.82 (0.71-0.95) |
| Sex |  |
| Female | 1.00 [Reference] |
| Male | 0.71 (0.67-0.76) |
| Race/Ethnicity |  |
| Non-Hispanic White | 1.00 [Reference] |
| Asian | 1.07 (0.85-1.34) |
| Hispanic | 0.89 (0.72-1.08) |
| Non-Hispanic Black | 1 (0.92-1.09) |
| Other/Unknown | 0.85 (0.72-0.99) |
| Insurance Plan |  |
| Commercial | 1.00 [Reference] |
| Medicaid | 1.26 (1.12-1.42) |
| Medicare | 1.15 (1.04-1.26) |
| Self-Pay/Other | 0.7 (0.6-0.82) |
| MyPennMedicine User |  |
| No | 1.00 [Reference] |
| Yes | 1.05 (0.96-1.14) |
| Marital Status |  |
| Married | 1.00 [Reference] |
| Unmarried | 1 (0.93-1.07) |
| Encounter Category |  |
| Return Patient Visit | 1.00 [Reference] |
| New Patient Visit | 0.44 (0.39-0.5) |
| Other/Unknown | - |
| Median household income |  |
| <$50,000 | 1.09 (1-1.2) |
| $50,000 to $100,000 | 1.00 [Reference] |
| ≥$100,000 | 1.12 (1.03-1.22) |
| Charlson Comorbidity Index score |  |
| <3 | 1.00 [Reference] |
| ≥3 | 0.6 (0.56-0.66) |
| Distance from Home to Place of Service |  |
| < 5 miles | 1.00 [Reference] |
| 5 to 15 miles | 1.11 (1.02-1.21) |
| ≥15 miles | 1.58 (1.45-1.74) |

### eTable 7. Adjusted odds ratios for COPD visits

| Characteristics | Adjusted Odds Ratio (95% CI) |
| --- | --- |
| Hospital |  |
| HUP | 1.00 [Reference] |
| CCH | 0.58 (0.46-0.73) |
| MCP | 0.55 (0.47-0.64) |
| PAH | 1.24 (1.11-1.39) |
| PPMC | 1.08 (0.98-1.19) |
| Age Range |  |
| < 40 years | 1.00 [Reference] |
| 40-64 years | 2.43 (1.28-4.61) |
| ≥65 years | 1.71 (0.9-3.26) |
| Sex |  |
| Female | 1.00 [Reference] |
| Male | 0.83 (0.77-0.89) |
| Race/Ethnicity |  |
| Non-Hispanic White | 1.00 [Reference] |
| Asian | 0.87 (0.6-1.26) |
| Hispanic | 1.72 (1.34-2.22) |
| Non-Hispanic Black | 1.25 (1.12-1.4) |
| Other/Unknown | 0.78 (0.63-0.96) |
| Insurance Plan |  |
| Commercial | 1.00 [Reference] |
| Medicaid | 1.44 (1.24-1.67) |
| Medicare | 1.23 (1.09-1.39) |
| Self-Pay/Other | 0.91 (0.76-1.1) |
| MyPennMedicine User |  |
| No | 1.00 [Reference] |
| Yes | 1.27 (1.16-1.39) |
| Marital Status |  |
| Married | 1.00 [Reference] |
| Unmarried | 1.11 (1.02-1.21) |
| Encounter Category |  |
| Return Patient Visit | 1.00 [Reference] |
| New Patient Visit | 0.31 (0.27-0.36) |
| Other/Unknown | - |
| Median household income |  |
| <$50,000 | 1.19 (1.06-1.33) |
| $50,000 to $100,000 | 1.00 [Reference] |
| ≥$100,000 | 0.99 (0.89-1.09) |
| Charlson Comorbidity Index score |  |
| <3 | 1.00 [Reference] |
| ≥3 | 0.65 (0.59-0.71) |
| Distance from Home to Place of Service |  |
| < 5 miles | 1.00 [Reference] |
| 5 to 15 miles | 0.89 (0.81-0.98) |
| ≥15 miles | 1.5 (1.35-1.68) |

### eTable 8. Adjusted odds ratios for CAD visits

| Characteristics | Adjusted Odds Ratio (95% CI) |
| --- | --- |
| Hospital |  |
| HUP | 1.00 [Reference] |
| CCH | 0.34 (0.28-0.41) |
| MCP | 0.47 (0.41-0.53) |
| PAH | 1.07 (0.99-1.17) |
| PPMC | 0.62 (0.57-0.67) |
| Age Range |  |
| < 40 years | 1.00 [Reference] |
| 40-64 years | 0.56 (0.44-0.72) |
| ≥65 years | 0.42 (0.33-0.54) |
| Sex |  |
| Female | 1.00 [Reference] |
| Male | 0.89 (0.83-0.95) |
| Race/Ethnicity |  |
| Non-Hispanic White | 1.00 [Reference] |
| Asian | 1.04 (0.9-1.21) |
| Hispanic | 0.91 (0.74-1.12) |
| Non-Hispanic Black | 1.14 (1.03-1.26) |
| Other/Unknown | 0.95 (0.83-1.09) |
| Insurance Plan |  |
| Commercial | 1.00 [Reference] |
| Medicaid | 0.99 (0.84-1.15) |
| Medicare | 1.16 (1.06-1.27) |
| Self-Pay/Other | 0.57 (0.49-0.67) |
| MyPennMedicine User |  |
| No | 1.00 [Reference] |
| Yes | 1.17 (1.08-1.28) |
| Marital Status |  |
| Married | 1.00 [Reference] |
| Unmarried | 1.02 (0.95-1.09) |
| Encounter Category |  |
| Return Patient Visit | 1.00 [Reference] |
| New Patient Visit | 0.35 (0.31-0.4) |
| Other/Unknown | - |
| Median household income |  |
| <$50,000 | 1.11 (1-1.24) |
| $50,000 to $100,000 | 1.00 [Reference] |
| ≥$100,000 | 1.02 (0.95-1.09) |
| Charlson Comorbidity Index score |  |
| <3 | 1.00 [Reference] |
| ≥3 | 0.73 (0.67-0.78) |
| Distance from Home to Place of Service |  |
| < 5 miles | 1.00 [Reference] |
| 5 to 15 miles | 1.18 (1.09-1.27) |
| ≥15 miles | 1.77 (1.63-1.92) |

### eTable 9. Adjusted odds ratios for GI visits

| Characteristics | Adjusted Odds Ratio (95% CI) |
| --- | --- |
| Hospital |  |
| HUP | 1.00 [Reference] |
| CCH | 0.2 (0.19-0.22) |
| MCP | 0.76 (0.73-0.78) |
| PAH | 1.04 (1.01-1.06) |
| PPMC | 0.89 (0.86-0.91) |
| Age Range |  |
| < 40 years | 1.00 [Reference] |
| 40-64 years | 0.82 (0.8-0.84) |
| ≥65 years | 0.6 (0.58-0.63) |
| Sex |  |
| Female | 1.00 [Reference] |
| Male | 0.81 (0.8-0.83) |
| Race/Ethnicity |  |
| Non-Hispanic White | 1.00 [Reference] |
| Asian | 0.83 (0.79-0.87) |
| Hispanic | 1.02 (0.98-1.06) |
| Non-Hispanic Black | 1.1 (1.07-1.13) |
| Other/Unknown | 0.88 (0.84-0.91) |
| Insurance Plan |  |
| Commercial | 1.00 [Reference] |
| Medicaid | 1.1 (1.06-1.13) |
| Medicare | 1.19 (1.15-1.22) |
| Self-Pay/Other | 0.64 (0.62-0.66) |
| MyPennMedicine User |  |
| No | 1.00 [Reference] |
| Yes | 1.45 (1.4-1.49) |
| Marital Status |  |
| Married | 1.00 [Reference] |
| Unmarried | 1.01 (1-1.03) |
| Encounter Category |  |
| Return Patient Visit | 1.00 [Reference] |
| New Patient Visit | 0.38 (0.37-0.39) |
| Other/Unknown | - |
| Median household income |  |
| <$50,000 | 1.1 (1.07-1.14) |
| $50,000 to $100,000 | 1.00 [Reference] |
| ≥$100,000 | 1 (0.98-1.02) |
| Charlson Comorbidity Index score |  |
| <3 | 1.00 [Reference] |
| ≥3 | 0.86 (0.84-0.88) |
| Distance from Home to Place of Service |  |
| < 5 miles | 1.00 [Reference] |
| 5 to 15 miles | 1.1 (1.07-1.12) |
| ≥15 miles | 1.44 (1.4-1.47) |
